# Effectiveness of BNT162b2 and ChAdOx1 vaccines against symptomatic COVID-19 among Healthcare Workers in Kuwait: A retrospective cohort study

**DOI:** 10.1101/2021.07.25.21261083

**Authors:** Walid Q. Alali, Lamiaa A. Ali, Mohammad AlSeaidan, Mohammad Al-Rashidi

## Abstract

**Background:** The COVID-19 BNT162b2 vaccination roll-out in Kuwait started on 24 December 2020 followed by ChAdOx1 on 3 February 2021. The study objectives were to assess the factors associated with vaccine coverage and determine vaccine effectiveness (VE) against SARS-CoV-2 infection in a healthcare worker (HCW) population.

**Methods:** This retrospective cohort study was conducted among HCW working at a public secondary hospital in Kuwait. The follow-up period was from 24 December 2020 to 15 June 2021. The primary outcomes were vaccine coverage and PCR-confirmed SARS-CoV-2 infection for the VE analysis. Data on new SARS-CoV-2 infections (with or without symptoms) during study period in addition to HCWs characteristics (sex, age, nationality, and occupation) were extracted from the hospital records. The vaccine coverage and PCR-confirmed SARS-CoV-2 infections were cross-tabulated by the HCWs characteristics. Furthermore, we used Cox regression to estimate time-to-infection hazard ratios in vaccinated (first and second dose) compared to unvaccinated HCWs. Only one ChAdOx1 dose was given during the study period.

**Results:** There were 3246 HCWs included in the analysis. The median age was 38 years (IQR = 33 - 44), 63.4% were females, 46.8% aged 31-40, and 82.3% were non-Kuwaitis. Overall, 82.1% of HCWs received at least one vaccine dose (50.4% received only one dose of ChAdOx1, 3.3% received one dose of BNT162b2, and 28.3% received two doses of BNT162b2). 17.9% of HCWs remained unvaccinated by the end of the study. A significantly lower vaccination coverage percentage was amongst female HCWs, younger age group (20 – 30 years old), and administrative/executive staff. Symptomatic SARS-CoV-2 PCR-confirmed infection prevalence was 7.3%. No asymptomatic infections were reported. The SARS-CoV-2 infection incidence rate was 126 per 100,000 person-days in the unvaccinated group; whereas, the incidence rates in the partially vaccinated groups (≥ 28 days after ChAdOx1 first dose) and (≥ 14 days after receiving BNT162b2 through receipt of second dose) were 31.4 and 10.9 per 100,000 person-days, respectively. In the fully vaccinated group (≥ 14 days after BNT162b2 second dose), the incidence rate was 6.3 per 100,000 person-days. The estimated adjusted vaccine effectiveness of fully vaccinated was 94.5% (95% confidence interval [CI] = 89.4%–97.2%). The VE of partially vaccinated for ChAdOx1 and BNT162b2 was 75.4% (95% CI = 67.2%–81.6%) and 91.4% (95% CI = 65.1%–97.9%), respectively.

**Conclusions:** Both BNT162b2 and ChAdOx1 vaccines prevented most symptomatic infections in this population across age groups, nationalities, and occupations. A significant proportion (17.9%) of HCWs were unvaccinated despite the vaccine accessibility. The findings complement other VE studies and demonstrate the vaccine benefit among HCWs.

## 1. Introduction

The two anti-COVID-19 vaccines (BNT162b2 mRNA [Pfizer-BioNTech] and ChAdOx1 nCoV-19 adenoviral [Oxford-AstraZeneca] have shown to be effective in preventing asymptomatic/symptomatic COVID-19, hospitalization, and death based on both clinical trials and population-level observational studies (Abu-Raddad, Chemaitelly, & Butt, 2021; Haas et al., 2021; Hall et al., 2021; Polack et al., 2020; Vasileiou et al., 2021; Voysey et al., 2021). Both vaccines have been authorized for use in Kuwait. Local BNT162b2 vaccination roll-out started on 24^th^ December 2020 followed by ChAdOx1 on 3^rd^ February 2021.

COVID-19 vaccine effectiveness under ‘real-world’ conditions are important to conduct not only in the general population but also in risk-specific groups such as healthcare workers (HCWs) due to their higher exposure rates to SARS-CoV-2. It is well-known that HCWs come in contact with patients directly and indirectly depending on their occupation. Therefore, they considered high risk group for SARS-CoV-2 infection and consequently at risk for disease complications. Recent studies have shown that vaccination reduce the rate of infection among HCWs (Amit, Regev-Yochay, Afek, Kreiss, & Leshem, 2021; Hall et al., 2021; Moustsen-Helms et al., 2021; Thompson et al., 2021). In the study from Israel, the incidence rate among BNT162b2 vaccinated HCWs was 3 cases per 10,000 person-days compared to 7.4 cases per 10,000 person-day in unvaccinated (Amit et al., 2021). Moreover, a study from United Kingdom (UK) revealed that in a single dose of BNT162b2 vaccine was 70% effective in reducing SARS-CoV-2 infections among HCWs 21 days after first dose and 85% after 7 days post second dose (Hall et al., 2021). Furthermore, a study from United States (US) on two mRNA vaccines (BNT162b2 and Moderna mRNA-1273) among HCWs, full immunization (≥14 days after second dose) was 90% effective against SARS-CoV-2 infections regardless of the symptom’s status and 80% effectiveness in partially immunized (≥14 days after first dose but before second dose).

While there are few studies that mostly evaluated impact of mRNA vaccines effectiveness in HCWs population; there is limited data on mRNA and non-mRNA vaccine effectiveness in HCWs population. Furthermore, there has been no published study on vaccine effectiveness among HCWs in Gulf Cooperation Council countries and most of Middle Eastern countries. Therefore, the objective of this retrospective cohort study was to assess the two vaccines (BNT162b2 and ChAdOx1) effectiveness against symptomatic SARS-CoV-2 infection and in relation to healthcare workers characteristics. We anticipate that the study findings will inform HCWs across the country, the region, and internationally of the importance of vaccination against COVID-19.

## 2. Methods

### 2.1. Study design and study population

The study was a retrospective cohort study among HCWs working at a public secondary hospital in Kuwait. The hospital is a 900-bed facility with multiple medical and surgical specialties including outpatient polyclinics. The original study population was 3673 HCWs (aged ≥ 20 years) working at this hospital. The study started on 24^th^ December 2020 (i.e., the day vaccine roll-out began in Kuwait). HCWs with PCR-confirmed SARS-CoV-2 before the start of the study were excluded. The study ended on 15^th^ June 2021. The two cohorts we followed were vaccinated and non-vaccinated HCWs. The main outcome of interest was symptomatic SARS-CoV-2 PCR-confirmed infections.

The study was approved by the Ministry of Health Standing Committee for the Coordination of Health and Medical Research (i.e., Ethics Committee), Kuwait City, Kuwait (Approval number: 1666/2021).

### 2.2. Data collection

Vaccination data were obtained from the hospital records. The hospital administration staff started collecting vaccination data on 10^th^ January 2021 from all HCWs. They regularly followed up with all hospital departments and requested specific vaccine-related data on HCWs including vaccination status (vaccinated or unvaccinated); vaccination date (for first and second dose); vaccine type (BNT162b2 or ChAdOx1); SARS-CoV-2 PCR-confirmed infections (symptomatic or asymptomatic) and infection date. Additionally, sociodemographic (sex, age, and nationality), occupation setting (e.g., outpatient, inpatient, intensive care), and staff occupation (e.g., doctor, nurse, pharmacist) were available in the hospital records. The sociodemographic and occupation data were matched with collected vaccination data using the civil identification number of the HCWs. The HCW testing for SARS-CoV-2 was voluntary on the basis of appearance of COVID-19 like symptoms or being a close-contact with COVID-19 positive case. A HCW was considered COVID-19 symptomatic if he/she had at least one typical disease symptoms such as fever, cough, or change in taste or smell. However, the vaccination records did not specify the range of symptoms but rather classified the infection as either symptomatic or asymptomatic.

The full data were extracted from the hospital records on 15^th^ June 2021. To avoid misclassification of exposure, HCWs with missing vaccination information (e.g., no vaccination date or vaccination type) or missing PCR testing information were excluded from the study. Furthermore, the analysis excluded 24 HCWs who had a documented SARS-CoV-2 PCR-confirmed infection prior to the study’s starting date and additional 403 HCWs who had missing or incomplete vaccination data or symptom data were also excluded. Hence, a total of 3246 HCWs eligible for this study.

### 2.3. Outcomes

The primary outcome for vaccine effectiveness analysis was the SARS-CoV-2 PCR-confirmed infection among unvaccinated or vaccinated at any time during the study (i.e., during the follow-up time) irrespective of symptom status. Infections were described as symptomatic if their symptom status was seven days before or seven days of their PCR positive test date.

The primary outcome for the vaccine coverage analysis was the vaccination status (first or second dose) by vaccine type. Healthcare workers vaccinated with ChAdOx1 had received only one dose by the end of study period. This was due to the delay in ChAdOx1 vaccine shipment to Kuwait that resulted in unavailability of the second dose for some HCWs who were vaccinated in February and March (12-week waiting period between two doses). The ChAdOx1 shipment arrived to Kuwait on 13^th^ June 2021; however, none of HCWs in this study received the second dose by 15^th^ June 2021.

### 2.4. Person-time at risk

The follow-up of all HCWs started on 24 December 2020, the day vaccine roll-out started in Kuwait. All HCWs had at least one day of follow-up as unvaccinated. For each HCW, the follow-up time (person-time at risk) ended at the earliest of the following events: occurrence of an outcome event (SARS-CoV-2 PCR-confirmed infection), vaccination (for unvaccinated), or end of study period.

### 2.5. Data analysis

Data were stratified by sociodemographic and occupation factors (i.e., covariates). These were: age group (20 -30, 31-40, 41 – 50, and >50), sex (male or female), nationality (Kuwaiti national and non-Kuwaiti resident), staff group (Administrative or Executive; Nursing or Health-care assistant; Doctor; Specialist Staff; Estates, Porters, or Security; and Pharmacist) and occupation settings (categorized into six groups: 1) office or laboratory, 2) hospital pharmacy, 3) outpatient including radiology, day ward, general practice, or renal dialysis unit, 4) inpatient ward, theatres, emergency department, maternity unit or labor ward, or ambulance, 5) intensive care, and 6) other (e.g., plaster and observational rooms).

For vaccine coverage analysis, we cross-tabulated three vaccination statuses as unvaccinated; vaccinated with one ChAdOx1 dose; and vaccinated with one or two doses of BNT162b2 with the study covariates. The relationship between vaccine coverage status and covariates were assessed via chi-square statistic using STATA software version 16.1 (College Station, Texas, USA). Furthermore, we also cross-tabulated SARS-CoV-2 PCR-confirmed infection by vaccination status and each level of the covariates. Similarly, chi-square statistic was used to assess relationships.

We used retrospective cohort study design to estimate the vaccine effectiveness in HCWs population after the first and second dose. For the purpose of vaccine effectiveness analysis, the HCWs were defined as unvaccinated (if they had not received any doses of either vaccine), fully vaccinated (if at least 14 days had passed since receiving the second dose of BNT162b2), and partial vaccination (if at least 28 days passed since receiving ChAdOx1 first dose or at least 14 days after receiving BNT162b2 first dose but before receiving second dose). The BNT162b2 13 person-days between receiving vaccine first dose and partial or full vaccination were considered excluded from the analysis as at-risk person-time because immunity was considered indeterminate. Similarly, the ChAdOx1 27 person-days after receiving vaccine first dose were excluded. Therefore, incidence rates were calculated for: unvaccinated, ≥ 28 days after receiving ChAdOx1 first dose, ≥ 14 days after receiving BNT162b2 first dose through receipt of second dose, and ≥ 14 days after BNT162b2 second dose. Hazard ratios were estimated using Cox proportional hazards model while accounting for time-varying vaccination status (i.e., receiving first and second dose) as described elsewhere (Zhang, Reinikainen, Adeleke, Pieterse, & Groothuis-Oudshoorn, 2018). Hazard ratios of partial vaccination person-days (≥ 28 days after receiving ChAdOx1 first dose; ≥14 days after receiving BNT162b2 first dose and before second dose) and to full vaccination person-days (≥14 days after BNT162b2 second dose) were calculated and compared to that in unvaccinated person-days. Vaccine effectiveness was calculated as 100% × (1-hazard ratio). An adjusted vaccine effectiveness model included the covariates individually (i.e., univariate models) and those were significant at P < 0.1 were included the multivariate model. All analyses were conducted in STATA statistical software.

## 3. Results

There were 3246 HCWs that met the inclusion criteria and included in the analysis. The median age of HCWs was 38 years (IQR = 33 - 44). Most of HCWs were females (63.4%), aged 31-40 (46.8%), non-Kuwaiti (82.3%), and worked in inpatient wards or ambulance settings (47.3%) as shown in Table 1. Furthermore, 61.2% of HCWs were nursing or health-care assistant staff.

**Table 1:**
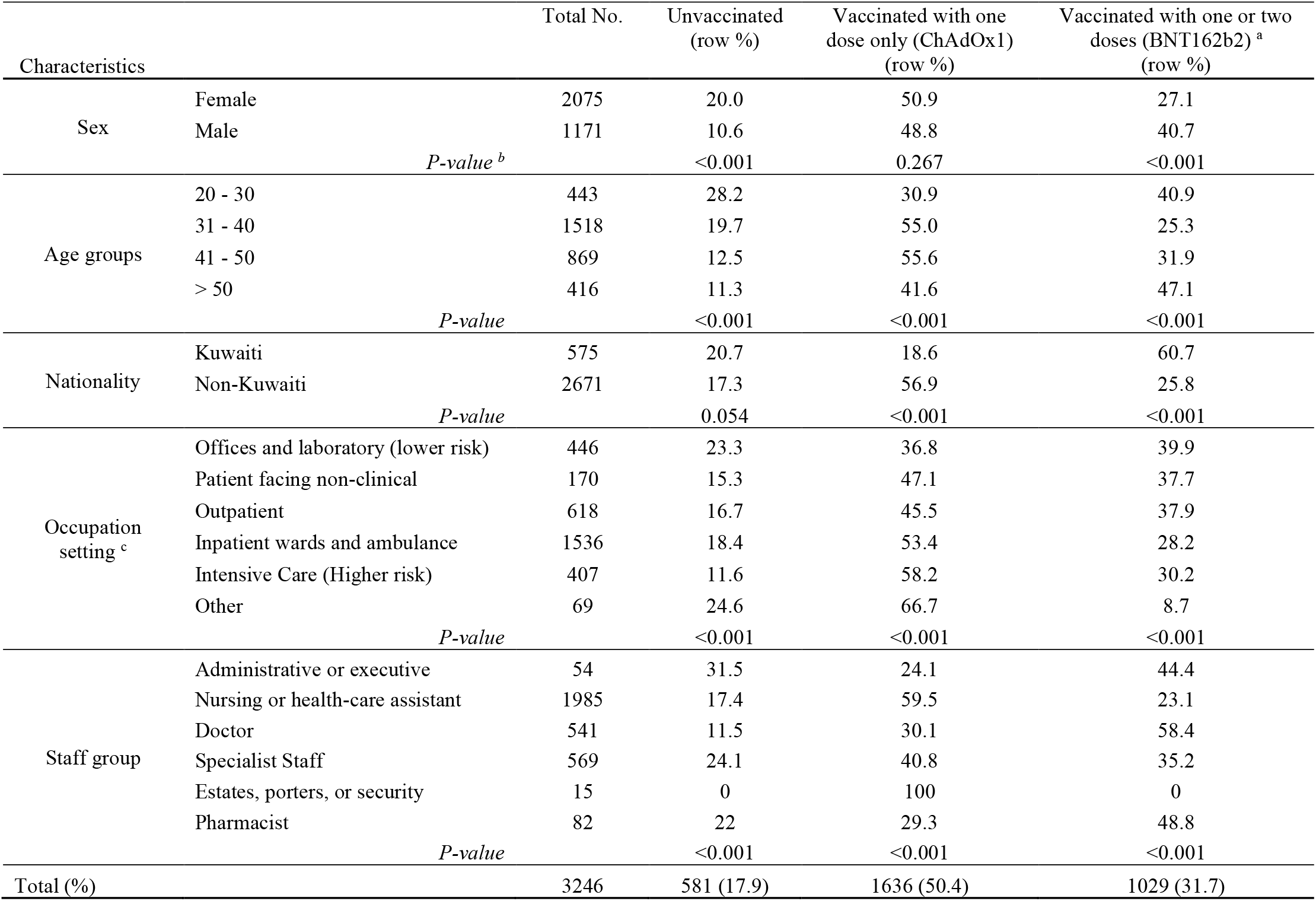
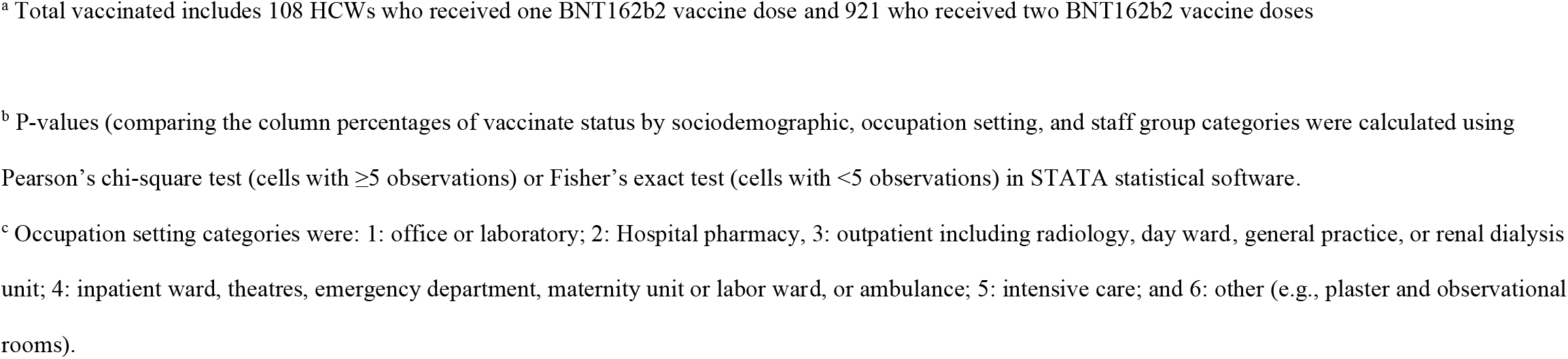
Characteristics of vaccinated and unvaccinated HCWs (n=3246) and factors associated with ChAdOx1 or BNT162b2 coverage at a major secondary hospital in Kuwait, December 24, 2020–June 15, 2021

| Characteristics |  | Total No. | Unvaccinated<br>(row %) | Vaccinated with one<br>dose only (ChAdOx1)<br>(row %) | Vaccinated with one or two<br>doses (BNT162b2) <sup>a</sup><br>(row %) |
| --- | --- | --- | --- | --- | --- |
| Sex | Female | 2075 | 20.0 | 50.9 | 27.1 |
|  | Male | 1171 | 10.6 | 48.8 | 40.7 |
| *P-value* <sup>b</sup> |  |  | <0.001 | 0.267 | <0.001 |
| Age groups | 20 - 30 | 443 | 28.2 | 30.9 | 40.9 |
|  | 31 - 40 | 1518 | 19.7 | 55.0 | 25.3 |
|  | 41 - 50 | 869 | 12.5 | 55.6 | 31.9 |
|  | > 50 | 416 | 11.3 | 41.6 | 47.1 |
| *P-value* |  |  | <0.001 | <0.001 | <0.001 |
| Nationality | Kuwaiti | 575 | 20.7 | 18.6 | 60.7 |
|  | Non-Kuwaiti | 2671 | 17.3 | 56.9 | 25.8 |
| *P-value* |  |  | 0.054 | <0.001 | <0.001 |
| Occupation<br>setting <sup>c</sup> | Offices and laboratory (lower risk) | 446 | 23.3 | 36.8 | 39.9 |
|  | Patient facing non-clinical | 170 | 15.3 | 47.1 | 37.7 |
|  | Outpatient | 618 | 16.7 | 45.5 | 37.9 |
|  | Inpatient wards and ambulance | 1536 | 18.4 | 53.4 | 28.2 |
|  | Intensive Care (Higher risk) | 407 | 11.6 | 58.2 | 30.2 |
|  | Other | 69 | 24.6 | 66.7 | 8.7 |
| *P-value* |  |  | <0.001 | <0.001 | <0.001 |
| Staff group | Administrative or executive | 54 | 31.5 | 24.1 | 44.4 |
|  | Nursing or health-care assistant | 1985 | 17.4 | 59.5 | 23.1 |
|  | Doctor | 541 | 11.5 | 30.1 | 58.4 |
|  | Specialist Staff | 569 | 24.1 | 40.8 | 35.2 |
|  | Estates, porters, or security | 15 | 0 | 100 | 0 |
|  | Pharmacist | 82 | 22 | 29.3 | 48.8 |
| *P-value* |  |  | <0.001 | <0.001 | <0.001 |
| Total (%) |  | 3246 | 581 (17.9) | 1636 (50.4) | 1029 (31.7) |

<sup>a</sup> Total vaccinated includes 108 HCWs who received one BNT162b2 vaccine dose and 921 who received two BNT162b2 vaccine doses
<sup>b</sup> P-values (comparing the column percentages of vaccinate status by sociodemographic, occupation setting, and staff group categories were calculated using
Pearson's chi-square test (cells with $\geq 5$ observations) or Fisher's exact test (cells with $< 5$ observations) in STATA statistical software.
<sup>c</sup> Occupation setting categories were: 1: office or laboratory; 2: Hospital pharmacy, 3: outpatient including radiology, day ward, general practice, or renal dialysis
unit; 4: inpatient ward, theatres, emergency department, maternity unit or labor ward, or ambulance; 5: intensive care; and 6: other (e.g., plaster and observational
rooms).

Overall, 82.1% of HCWs received at least one vaccine dose while 17.9% of HCWs remained unvaccinated by the end of the study. Interestingly, about half of the HCWs (50.4%) received only one dose of ChAdOx1 vaccine; whereas 3.3% received one dose of BNT162b2 vaccine and 28.3% received two doses of BNT162b2 vaccine. Those who received only one dose of BNT162b2 (3.3%) did not received their second dose because they were SARS-CoV-2 infected after the first dose (only two HCWs) or the study ended before they received it.

The percentage of HCWs classified as partially vaccinated (i.e., ≥ 28 days after receiving one dose of ChAdOx1 or ≥ 14 days after receiving BNT162b2 first dose through receipt of second dose) was 50.2% and 2.8%, respectively. However, the percentage of HCWs classified as fully vaccinated (≥ 14 days after BNT162b2 second dose) was 28.2%.

The characteristics of unvaccinated and vaccinated HCWs by the two types of vaccine are shown in Table 1. Twenty percent of females were unvaccinated compared to 10.6% of males (P <0.001). For age groups, 28.2% of HCWs aged 20-30 were unvaccinated, significantly higher than other age groups (P <0.001); whereas, within those received one or two doses of BNT162b2 vaccine, the percentage of vaccinated HCWs in age groups (20 – 30 and > 50) was higher than that in other age groups (P <0.001). Interestingly, the percentages of unvaccinated Kuwaitis (20.7%) and non-Kuwaitis (17.3%) HCWs were not significantly different (P = 0.054); however, within those received one or two doses of BNT162b2, 60.7% of Kuwaiti HCWs were vaccinated compared to 25.8% for non-Kuwaitis (P <0.001). Among the different occupation settings, 23.3% of HCWs who worked in office or laboratories were unvaccinated, significantly higher compared to the remaining settings (P <0.001) except for ‘other’. As for the HCW staff groups, 31.5% of administrative or executive staff were unvaccinated, significantly higher than other groups (P <0.001). In addition, 58.4% of doctors received one or two doses of BNT162b2 vaccine significantly higher than other staff groups (P <0.001).

SARS-CoV-2 PCR-confirmed infection prevalence with reported symptoms was 7.3% (237/3246) during the study period. There were two additional SARS-CoV-2 PCR-confirmed HCW infections with missing symptomatic status; hence, they were excluded from the analysis. Therefore, all the 237 SARS-CoV-2 PCR-confirmed infection were classified as symptomatic. As shown in Table 2, the infection prevalence was significantly higher among unvaccinated female HCWs (20.8%) compared to 7.1% in those vaccinated with ChAdOx1 and 1.96% in those vaccinated with one or two doses of BNT162b2. Similar findings were observed among male HCWs. SARS-CoV-2 PCR-confirmed prevalence in the different age-groups and by nationality were significantly higher in unvaccinated compared to the vaccinated groups. Furthermore, the infection prevalence was significantly higher across the unvaccinated occupation settings compared to those in vaccinated occupation settings except for “other” where the differences were not significant (P = 0.508). The infection prevalence in staff groups were also significantly higher in unvaccinated compared to vaccinated except for pharmacists (P = 0.866) (Table 2).

**Table 2:**
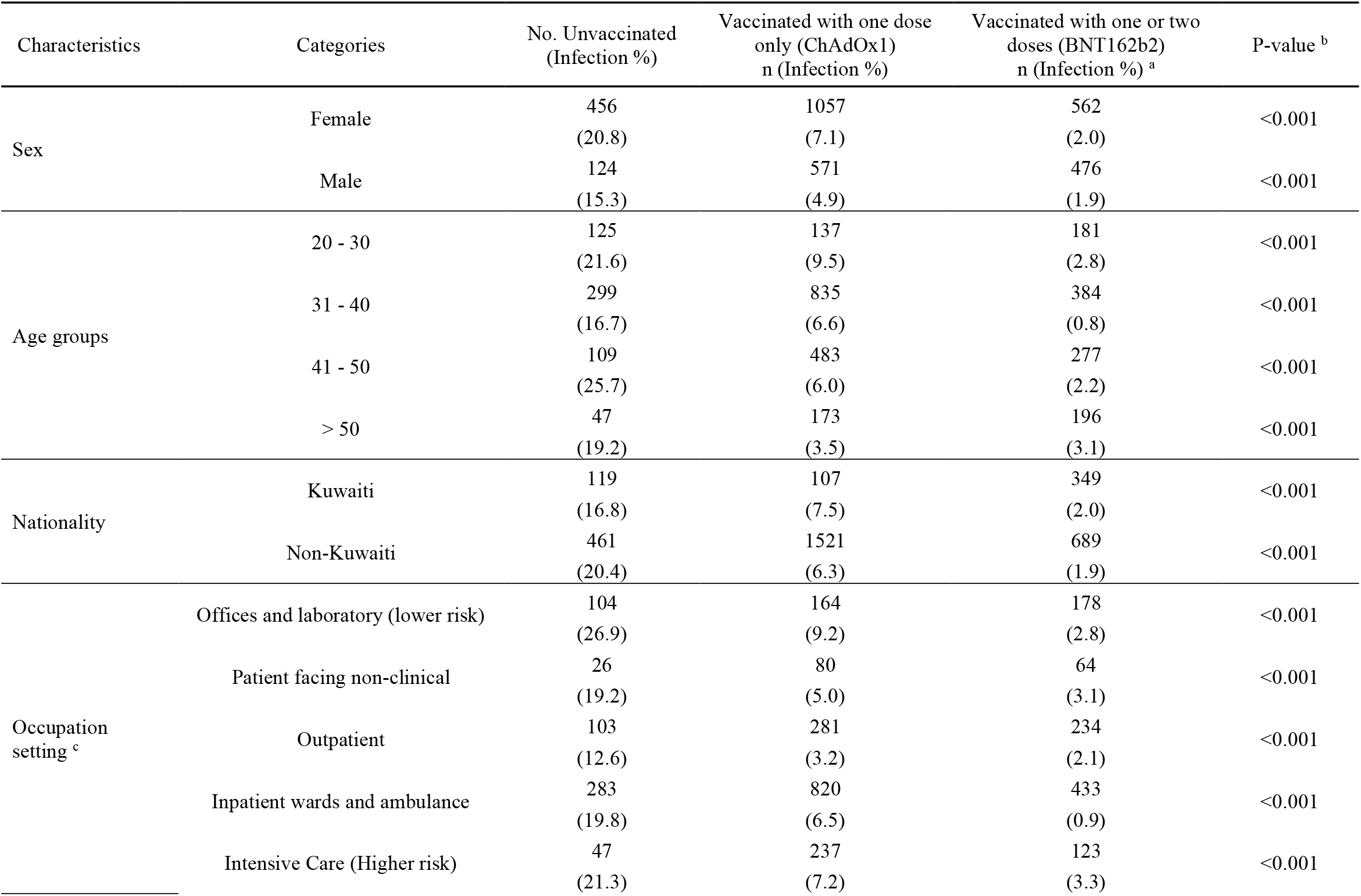
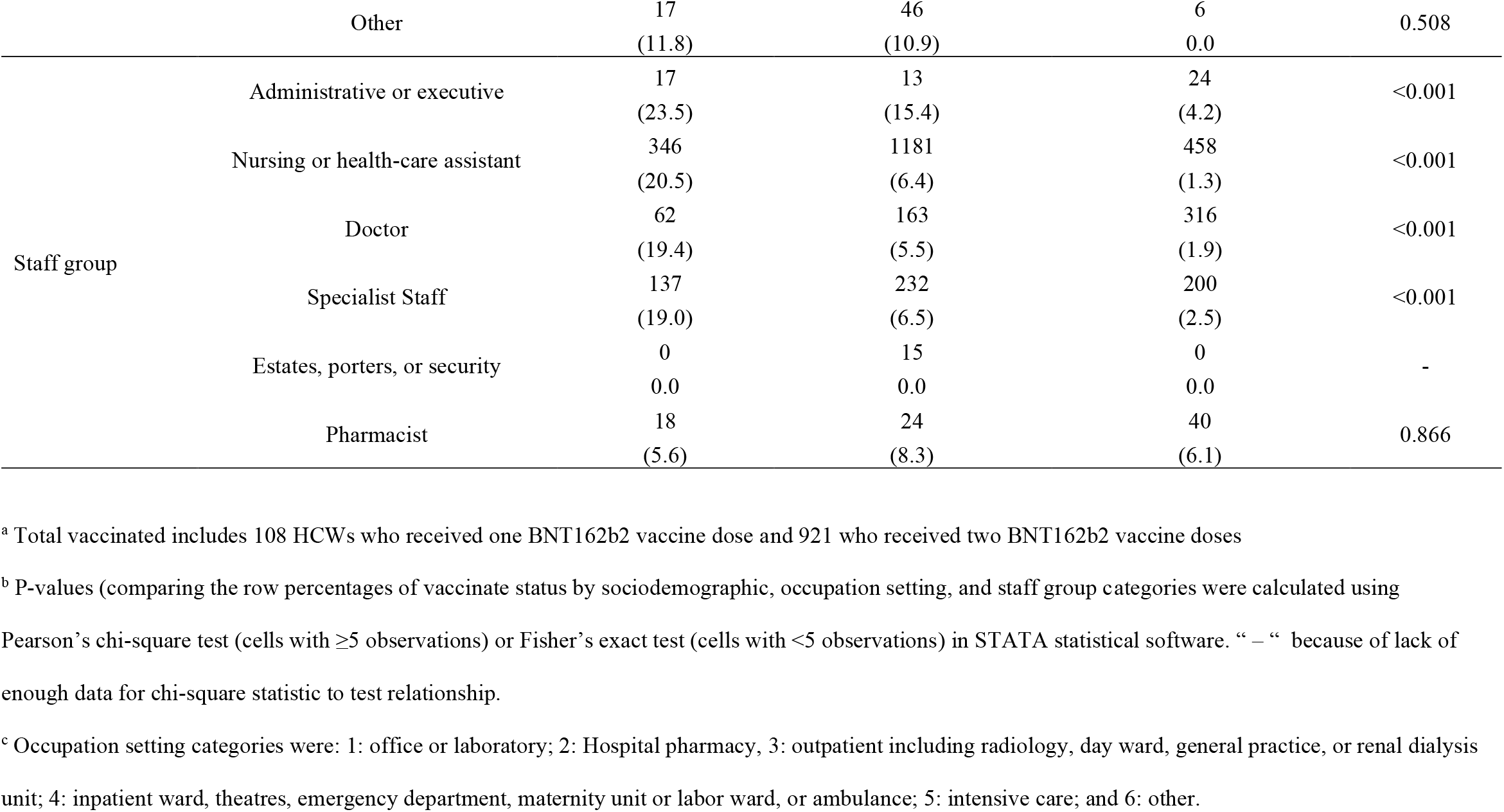
Prevalence of SARS-CoV-2 PCR–confirmed infections in HCWs (n=3246) received BNT162b2 or ChAdOx1 COVID-19 vaccines at a major secondary hospital in Kuwait, December 24, 2020–June 15, 2021

There were 114 SARS-CoV-2 infection during the 90,484 person-days of follow-up in the unvaccinated group, an incidence rate of 126 per 100,000 person-days (Table 3). In the partially vaccinated group, ≥ 28 days after ChAdOx1 first dose, there were 87 infections (incidence rate of 31.4 per 100,000 person-days). Moreover, in the partially vaccinated group (≥ 14 days after receiving BNT162b2 vaccine through receipt of second dose), there were two infections (incidence rate of 10.9 per 100,000 person-days). In the fully vaccinated group (≥ 14 days after BNT162b2 second dose) there were 10 infections (incidence rate of 6.3 per 100,000 person-days).

**Table 3.** Effectiveness of ChAdOx1 and BNT162b2 COVID-19 vaccines against SARS-CoV-2 symptomatic infection among HCWs (n= 3246) at a major secondary hospital in Kuwait, December 24, 2020–June 15, 2021

| COVID-19 vaccination status | Total person time (days) | Number of PCR positives | Incidence rate per 100,000 person-days | Unadjusted vaccine effectiveness % (95% CI) <sup>a</sup> | Adjusted vaccine effectiveness % (95% CI) <sup>a, b</sup> |
| --- | --- | --- | --- | --- | --- |
| Unvaccinated | 90,367 | 114 | 126.2 | Reference | Reference |
| Partially vaccinated |  |  |  |  |  |
| ≥ 28 days after receiving ChAdOx1 first dose only <sup>c</sup> | 159,423 | 87 | 54.6 | 75.5 (67.6 - 81.5) | 75.4 (67.2 - 81.6) |
| ≥ 14 days after receiving BNT162b2 first dose through receipt of second dose | 7,196 | 2 | 27.8 | 91.6 (65.9 - 97.9) | 91.4 (65.1 - 97.9) |
| Fully vaccinated |  |  |  |  |  |
| ≥ 7 days after BNT162b2 second dose | 90,015 | 12 | 13.3 | 95.1 (90.6 - 97.4) | 94.5 (89.4 - 97.2) |

<sup>a</sup> Vaccine effectiveness was estimated using a Cox proportional hazards model accounting for time-varying immunization status in STATA statistical software.
<sup>b</sup> Hazard ratio is adjusted for age, sex, and nationality
<sup>c</sup> Participants received first dose of ChAdOx1 but had not received second dose by the end of the study period.

The estimated adjusted vaccine effectiveness of fully vaccinated was 94.5% (95% confidence interval [CI] = 89.4%–97.2%). The vaccine effectiveness of partially vaccinated for ChAdOx1 (≥ 28 days after one dose) was 75.4% (95% CI = 67.2%–81.6%) and for one dose BNT162b2 (≥ 14 days through receipt of second dose) was 91.4% (95% CI = 65.1%–97.9%) (Table 3). The individual covariates (sex, age group, nationality, occupation setting, staff group) were significant predictors; hence, included in the multivariate model. However, these covariates were not significant (P > 0.05) in the adjusted vaccine effectiveness multivariate model. We kept the sociodemographic variables (sex, age group, and nationality) in the adjusted model and compared the change between unadjusted and adjusted models. The change in vaccine effectiveness point estimates were <1% between unadjusted and adjusted model.

## 4. Discussion

This retrospective cohort study was conducted between 24 December 2020 and 15 June 2021 (i.e., 173 days) at a secondary hospital in Kuwait shows that full vaccination (i.e., immunization) via BNT162b2 is highly effective against symptomatic COVID-19 among this HCW population. Furthermore, ChAdOx1 one dose was relatively effective (Table 3).

Vaccine coverage with at least one dose among HCWs after 173 days (about 5.8 months) was 82.1% of HCWs including 28.3% who received two doses. However, still there were 17.9% of HCWs unvaccinated by the end of the study which is a concern. Healthcare workers have been given the priority for vaccination in Kuwait as most countries; therefore, efforts are needed to better understand reasons for vaccine hesitancy in this high-risk exposure group. Other studies have reported that most HCWs were vaccinated with at least one dose within two to three months of vaccine roll-out (i.e., 90% in UK; 79% in Israel; and 75% in USA) (Amit et al., 2021; Hall et al., 2021; Thompson et al., 2021).

There were significant differences in vaccine coverage by demographics, occupation setting, and staff group. Similar differences by factors such as age, sex, ethnicity, and occupation have been reported in other studies (Curtis et al., 2021; Galanis, Vraka, Fragkou, Bilali, & Kaitelidou, 2020; Martin et al., 2021). The differences in vaccine coverage we reported among this population highlights the importance of equitable vaccination program to all HCWs in Kuwait.

Vaccine effectiveness of full vaccination with two doses of BNT162b2 vaccine was 94.5% (95% CI = 89.4%–97.2%) against symptomatic PCR-confirmed SARS-CoV-2 infection, whereas it was 75.4% (95% CI = 67.2%–81.6%) for ChAdOx1 single dose. These findings are consistent with those from other population-level studies that estimated vaccine effectiveness against SARS-CoV-2 infection (symptomatic and/or asymptomatic) among HCWs (Amit et al., 2021; Hall et al., 2021; Moustsen-Helms et al., 2021; Thompson et al., 2021) and those from vaccine phase III trials (Knoll & Wonodi, 2021; Polack et al., 2020). For instance, in a study from the US CDC, the authors reported that vaccine effectiveness against SARS-CoV-2 infection among HCWs with full immunization (≥ 14 days after BNT162b2 second dose) was 90% (95% CI = 68% – 97%) (Thompson et al., 2021). In other studies on HCWs, BNT162b2 vaccine effectiveness against SARS-CoV-2 infection (≥ 7 day post second dose) was 85% (95% CI = 74% – 96%) in the UK (Hall et al., 2021), and it was 85% (95% CI = 71% - 92%) 15 – 24 days after second dose in Israel (Amit et al., 2021). The main difference between these studies and ours is that HCWs were regularly tested for SARS-CoV-2 infection (active surveillance) while in our study it was based on reported of PCR-confirmed infection by HCWs to the hospital management (passive surveillance). Nonetheless, it is mandatory for HCWs to report if they were in close contact with a positive case or have tested positive via nasopharyngeal PCR.

Our findings highlight the effectiveness of vaccine in reducing the risk of symptomatic infection among HCWs across sociodemographic factors (sex, age, and nationality) and across occupation setting and staff group. Importantly, reducing infection rate among HCWs via vaccination is critical to protect their health and lower the transmission risk to their contacts (coworkers and patients) as well as to the public (Buitrago-Garcia et al., 2020).

The partial vaccination (≥ 28 days after ChAdOx1 one dose) provided about 75% protection. This similar with to the Phase III trial results (Hyams et al.; Knoll & Wonodi, 2021). However, to best of our knowledge, there is no estimate on vaccine effectiveness of ChAdOx1 in a HCW population. Furthermore, partial vaccination (≥14 days after first dose but before second dose) also provided high level of protection from infection in HCWs in this study; however, this limited by the relatively short at-risk person-time. Recent studies showed that partial vaccination among HCWs in the U.S. (≥ 21 days after BNT162b2 first dose) had vaccine effectiveness of 80% (95% CI = 59% – 90%) and 72% (95% CI = 58%–86%) in a study from UK (Hall et al., 2021; Thompson et al., 2021). Both studies were based on regular SARS-CoV-2 testing program. Moreover, a study from Israel reported 60% (95% CI = 38%–74%) one dose BNT162b2 vaccine effectiveness against confirmed SARS-CoV-2 infection based on hospital records (passive reporting) (Amit et al., 2021). It is worth mentioning that SARS-CoV-2 B.1.1.7 (Alpha) variant was detected in Kuwait in January 2021 and could have been the dominant variant during the study period; however, there are no available data in Kuwait to determine the percentage and distribution of infections with this variant. Moreover, the B.1.617.2 (Delta) variant was reported in Kuwait in late June 2021 (after the study ended); hence, it did not confound the effectiveness results.

This study has several limitations. First, the study was based on one public secondary hospital and might not be generalizable to HCWs in other public hospitals in Kuwait. However, this hospital is one of the major healthcare facilities in Kuwait and serves over a quarter of the country’s population. Second, the identification of HCWs SARS-CoV-2 PCR-confirmed infections was based on passive reporting to the hospital management due to lack of active laboratory surveillance. However, it was/is required by all HCWs to report PCR-confirmed infections to their upper management within each hospital’s department. Furthermore, underreporting PCR-confirmed infections might underestimate the ‘actual’ number of infections regardless of vaccination status; if this disproportionately impacted those who were unvaccinated compared to those who were vaccinated, this could overestimate vaccine effectiveness. Third, vaccine effectiveness estimates for partial immunity (≥ 14 days after BNT162b2 first dose through receipt of second dose) had wide confidence intervals which likely due to the low number of PCR-confirmed infections reported.

In conclusion, vaccine effectiveness of both BNT162b2 and ChAdOx1 COVID-19 vaccines in HCW under ‘real-world’ conditions demonstrated that vaccine is effective in preventing most symptomatic infection across age groups, nationalities, occupation setting, and staff groups. A significant proportion (17.9%) of HCWs were unvaccinated despite the vaccine accessibility. Efforts are needed to better understand reasons for HCW vaccine hesitancy. Although vaccination is highly effective against infection, hospitalization, and mortality as shown in other studies, it is important for HCWs to continue to exercise physical distancing, wear personal protective equipment while in contact with patients, and follow other infection control and prevention measures.

## Data Availability

Data are available based on request to the corresponding author

## Notes

### Competing Interest Statement

The authors have declared no competing interest.

### Funding Statement

No funding was received

